# Asymptomatic and mildly symptomatic influenza virus infections by season -- Case-ascertained household transmission studies, United States, 2017-2023

**DOI:** 10.1101/2024.07.17.24310569

**Authors:** Jessica E. Biddle, Huong Q. Nguyen, H. Keipp Talbot, Melissa A. Rolfes, Matthew Biggerstaff, Sheroi Johnson, Carrie Reed, Edward A. Belongia, Carlos G. Grijalva, Alexandra M. Mellis

**Affiliations:** Centers for Disease Control and Prevention, Atlanta, Georgia; Marshfield Clinic Research Institute, Marshfield, Wisconsin; Vanderbilt University Medical Center, Nashville, Tennessee

## Abstract

Asymptomatic influenza virus infection occurs but may vary by factors such as age, influenza vaccination status, or influenza season. We examined the frequency of influenza virus infection and associated symptoms using data from two case-ascertained household transmission studies (conducted from 2017—2023) with prospective, systematic collection of respiratory specimens and symptoms. From the 426 influenza virus infected household contacts that met our inclusion criteria, 8% were asymptomatic, 6% had non-respiratory symptoms, 23% had acute respiratory symptoms, and 62% had influenza-like illness symptoms. Understanding the prevalence of asymptomatic and mildly symptomatic influenza cases is important for implementing effective influenza prevention strategies and enhancing the effectiveness of symptom-based surveillance systems.

## Introduction

Influenza virus infections cause a range of health outcomes, spanning from asymptomatic infection to severe illness including hospitalizations and death. The COVID-19 pandemic has underscored the significance of asymptomatic transmission of respiratory pathogens, prompting further research to understand the frequency of asymptomatic infection with influenza virus and its implications on transmission. However, studying asymptomatic and mildly symptomatic influenza virus infections is challenging because of the need for prospective and active follow-up [1]. Even with prospective follow-up, estimates of the asymptomatic fraction have been heterogeneous, varying by experimental setting, study design, and case definitions for describing infections as asymptomatic [2-6].

The occurrence of asymptomatic influenza virus infection may also vary by age, vaccination status, and infecting virus type and subtype, but this heterogeneity is not well described. Understanding how these factors can influence asymptomatic influenza is crucial for developing effective public health strategies, as they can significantly impact the transmission dynamics and overall burden of influenza in populations. The objective of this analysis was to examine the proportion of asymptomatic influenza virus infections among household contacts and explore the asymptomatic fraction by individual-level factors using recent case-ascertained household studies of influenza that systematically collected daily respiratory specimens and symptoms.

## Methods

Two case-ascertained household transmission studies were conducted in Tennessee and Wisconsin across five influenza seasons from 2017–2023. Index cases with laboratory-confirmed influenza virus infection were enrolled within 7 days of symptom onset. At enrollment, all participants self-reported demographics, household characteristics, medical history, symptoms in the week prior to enrollment, and influenza vaccine receipt for the current season. Both index cases and their household contacts were eligible to enroll. Household contacts were defined as those who (1) slept in the same household as the index case for at least half the preceding month, (2) who were present for at least one night in the week before enrollment, and (3) planned to continue residing in the household during the follow-up period. Enrolled household members had nasal swabs collected daily, for 5 days (2017-2020 in Wisconsin), 7 days (2017-2020 in Tennessee), or 10 days (2021-2023 at both sites). During these sampling periods, participants also completed symptom diaries daily. Nasal swabs were tested for influenza viruses using reverse transcriptase polymerase chain reaction (RT-qPCR). Influenza positive specimens were subtyped according to methods previously described⍰⍰. Participants reported presence or absence of the following signs and symptoms: fever, cough, runny nose, nasal congestion, fatigue, wheezing, shortness of breath, and sore throat on each daily symptom diary. Protocols and procedures were reviewed and approved by the institutional review boards (IRB) at the Marshfield Clinic Research Institute and Vanderbilt University from 2017-2020 and a single IRB at Vanderbilt University from 2021-2023.

Analyses were restricted to household contacts who tested positive for influenza. Contacts were excluded if they had less than 3 lab results or daily diaries, or if their first specimen was taken more than 7 days from the index’s onset date. Analyses were also restricted to up to 7 days of follow-up. As a sensitivity analysis, we also restricted to 5 days of follow-up, which was the minimum duration of symptom surveillance collected at any site during any season. For analyses, we used data from all available daily diaries and implemented 3 definitions to classify the infection: (1) asymptomatic vs symptomatic, (2) without acute respiratory illness (ARI) vs with ARI, and (3) without influenza-like illness (ILI) vs with ILI. Being symptomatic was defined as having at least one of the following signs or symptoms: fever, cough, runny nose, nasal congestion, fatigue, wheezing, shortness of breath, or sore throat. ARI was defined as having two or more of the following signs or symptoms: fever, cough, runny nose, nasal congestion, or sore throat [8]. ILI was defined as having fever and cough or fever and sore throat. For descriptive purposes, participants were classified into 4 mutually exclusive groups: (1) asymptomatic overall, (2) symptomatic, non-ARI, (3) ARI but not ILI, and (4) ILI.

Three multivariable, robust logistic regression models were conducted for each symptom definition to estimate the proportion of influenza positive household contacts who were asymptomatic, did not develop ARI, and did not develop ILI. All regression models accounted for household clustering. Each of the three regression models were further stratified by age group, by season, and by vaccination status. Regression models stratified by age group were adjusted for season and vaccination status. Regression models stratified by season were adjusted for age group and vaccination status. And finally, regression models stratified by vaccination status were adjusted for age group and season. These regression models were used to generate predicted probabilities of the fraction of infected contacts with each symptom presentation, which are presented by age, by season, and by vaccination status.

## Results

Four hundred and twenty-six household contacts tested positive for influenza and met our criteria for inclusion. Fifty-four percent of included contacts were from Tennessee, 50% were male, 49% were aged <18 years, 73% were white, non-Hispanic, and 60% were unvaccinated against influenza during the season they were enrolled (Supplemental Table 1). Influenza viruses detected in the infected contacts varied by season (Supplemental Table 1).

From the 426 infected household contacts, 36 (8%) were asymptomatic, 27 (6%) had symptoms but not-ARI, 98 (23%) had ARI but not ILI, and 265 (62%) had ILI during the follow-up period (Figure 1). The proportion of asymptomatic or mildly symptomatic (without ARI) contacts was highest among adult participants (ages ≥18 years) and those who were vaccinated (Figure 1). Additionally, the proportion of asymptomatic or mildly symptomatic contacts was highest in seasons after the COVID-19 pandemic started compared to the pre-pandemic seasons (Figure 1).

**Figure 1.**
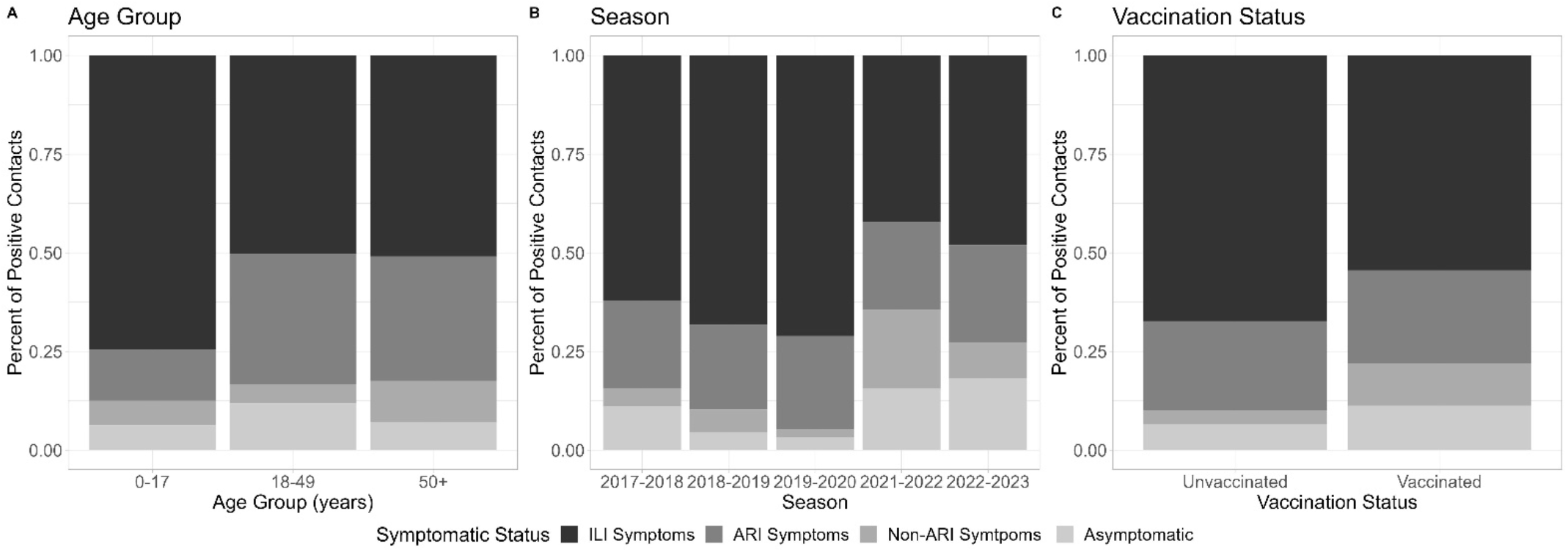
Symptom status of influenza positive contacts by age group, season, and vaccination status, United States, 2017 - 2023 Symptomatic status was defined using the following symptom definitions (1) **Influenza-like Illness (ILI) Symptoms:** Fever + (Cough OR Sore Throat), (2) **Acute Respiratory Illness (ARI) Symptoms:** 2 or more of the following symptoms: Fever, Cough, Runny Nose, Nasal Congestion, Sore Throat, and (3) **Non-ARI Symptoms:** At least one of the following symptoms: Fever, Cough, Runny Nose, Nasal Congestion, Fatigue, Wheezing, Shortness of Breath, Sore Throat. From the 426 included infected household contacts 36 (8%) were completely asymptomatic, 27 (6%) had non-ARI symptoms, 98 (23%) had ARI symptoms and 265 (62%) had ILI symptoms.

We completed three robust multivariable logistic regression models accounting for age, season, and vaccination against each binary case definition for asymptomatic and mildly symptomatic infections. The prevalence of asymptomatic infections across age groups varied: 8% for ages 0–17, 12% for ages 18–49, and 5% for ages ≥50 years. Similarly, absence of ARI was observed in 16% of contacts for ages 0–17, 17% for ages 18–49, and 13% for ages ≥50 years, while absence of ILI occurred in 30% for ages 0– 17, 54% for ages 18–49, and 48% for ages ≥50 years (Table 1). Notably, the prevalence of asymptomatic infections was highest in the 2021-2022 and 2022-2023 seasons compared to pre-pandemic seasons. Additionally, adjusting for age group and season, 12% of vaccinated contacts and 5% of unvaccinated contacts were asymptomatic. Vaccinated contacts also showed lower percentages of ARI (9% vs 25%) and ILI (36% vs. 52%) compared to unvaccinated contacts (Table 1). Trends by age group, season, and vaccination status remained the same when assessments were restricted to the first 5 days of follow-up (Supplemental Table 2).

**Table 1.** Characteristics of the cohort & frequencies of asymptomatic infections, United States, 2017-2023.

|  | Influenza positive contacts | Asymptomatic influenza positive contacts | Modeled Asymptomatic Infections* | Contacts without ARI | Modeled Infections without ARI* | Contacts without ILI | Modeled Infections without ILI* |
| --- | --- | --- | --- | --- | --- | --- | --- |
|  | N | N | % (CI) | N | % (CI) | N | % (CI) |
| **Age Group** |  |  |  |  |  |  |  |
| 0 – 17 years | 208 | 13 | 8 (4-14) | 26 | 16 (11-24) | 53 | 30 (23-39) |
| 18 – 49 years | 161 | 19 | 12 (7-20) | 27 | 17 (11-26) | 80 | 54 (44-63) |
| 50+ years | 57 | 4 | 5 (2-15) | 10 | 13 (7 – 23) | 28 | 48 (35-61) |
| **Season** |  |  |  |  |  |  |  |
| 2017 – 2018 | 45 | 5 | 10 (4-23) | 7 | 15 (7-29) | 17 | 38 (24-55) |
| 2018 – 2019 | 107 | 5 | 4 (2- 9) | 11 | 8 (4-16) | 34 | 34 (24-45) |
| 2019 – 2020 | 152 | 5 | 3 (1-9) | 8 | 5 (2-11) | 44 | 34 (26-43) |
| 2021 – 2022 | 45 | 7 | 15 (7-30) | 16 | 36 (21-53) | 26 | 59 (40-75) |
| 2022 – 2023 | 77 | 14 | 17 (9-31) | 21 | 30 (18-46) | 40 | 55 (42-68) |
| **Influenza Vaccination Status** |  |  |  |  |  |  |  |
| Vaccinated | 169 | 19 | 12 (8-18) | 37 | 25 (17-34) | 77 | 52 (42-61) |
| Unvaccinated | 257 | 17 | 5 (3-10) | 26 | 9 (6-14) | 84 | 36 (29-44) |

\*A robust logistic regression model accounting for household clustering was conducted to estimate the proportion of influenza positive household contacts that were asymptomatic, without ARI, and without ILI. Adjusted marginal estimates and 95% confidence intervals are shown as percentages. Age group was adjusted for season and vaccination status. Season was adjusted for age group and vaccination status. Vaccination status was adjusted for age group and season.

## Discussion

In this study, the proportion of asymptomatic infection among individuals with laboratory-confirmed influenza was 8% overall. Mildly symptomatic infections, characterized by the absence of ARI, accounted for 6% of cases, while mildly symptomatic infections, characterized by the presence of ARI but not ILI, comprised 23% of cases. Consistent with previous studies, we found that asymptomatic influenza virus infections occur and further show that the asymptomatic fraction, as we estimated using a household transmission study, varies by season and, possibly, influenza vaccination.

Understanding the fraction of individuals with influenza who are asymptomatic or have only mild symptoms has implications for both the prevention of influenza through transmission control and the ability to detect influenza through symptom-based surveillance systems, particularly as these infections may be more common than previously thought, underscoring the significance of our updated evidence. As highlighted by a review by Montgomery et al., if a significant proportion of influenza transmission occurs through individuals with mild or no symptoms, prevention strategies beyond symptom-based case identification, such as vaccination, gain heightened importance [1]. In addition, if the fraction of individuals who meet standard case definitions for influenza changes over time or varies age groups, this has implications for continued ability to detect influenza or estimate total illnesses or infections.

The present study showed a trend of increased asymptomatic and mildly symptomatic influenza virus infections during the post COVID-19 pandemic seasons. Following historically low levels of influenza activity in 2020, which persisted into 2021 [9], there was an increase in the risk of influenza virus infection among household contacts in the 2021-2022 season compared to pre-pandemic seasons [7]. Further research is needed to understand the factors that are driving the increase in infections post-pandemic. We hypothesize the increase in infections could be due to increased susceptibility among individuals without recent exposure. We further see that during the 2021-2022 season, influenza virus infections in this study may have been more mild or were asymptomatic.

Previous studies have examined asymptomatic fraction by age and vaccination status. Consistent with our findings, previous research has shown asymptomatic influenza virus infection among children is less common compared to adults [10]. Findings from the household cohort study out of South Africa further underscores this age-related pattern, showing that the rates of influenza virus infection and symptomatic illnesses were highest in children <5 years and decreased with increasing age [3]. Studies exploring the association of influenza vaccination and asymptomatic fractions within households are lacking. However, one previous study examining the protective effect of influenza vaccination against symptomatic infection in healthcare workers demonstrated that vaccinated health care workers had a higher proportion of asymptomatic influenza virus infection compared to unvaccinated health care workers which is consistent with our present findings [11]. Further research with larger sample sizes should explore the association between vaccination status and progression of symptoms.

There are several limitations to our study. First, our study recruited index cases who sought medical care for their illness. Thus, our analysis would not have included household contacts exposed to index cases with milder or asymptomatic presentations that would likely have not sought medical care. Second, serological data, which have been previously used to generate estimates of asymptomatic infections, were not included in this study and therefore we cannot supplement the ascertainment of asymptomatic cases with those data. Additionally, we were unable to stratify data into finer-resolved age groups or influenza types/subtypes due to sample size constraints across seasons, limiting the ability to discern more age or influenza-type specific patterns of asymptomatic or mildly symptomatic infections. This analysis was restricted to 7 days of active follow-up (median of 9 days of follow up after first illness in the household) and some individuals who were classified as asymptomatic may have developed symptoms later. Finally, we collected data exclusively from households in two US states and therefore our results may not be generalizable to broader populations. Looking forward, future research efforts should consider inclusion of more diverse demographic and geographic settings to ensure a comprehensive understanding of asymptomatic influenza virus infections.

These results underscore the value of household transmission studies with close follow-up of exposed household members to better understand the spectrum of symptoms seen in influenza virus infection, including the rate of asymptomatic infection among individuals with influenza.

## Data Availability

The data that support the findings of this study are available on request from the corresponding author. The data are not publicly available due to privacy or ethical restrictions.

## Acknowledgements

The authors would like to thank Alicia Fry, Scott Blackwell, Emma Pedigree Cannon, Holly Morse, Megan Eluhu, Emily Jookar, Christina Khouri, Robert Lyons, Carleigh Frazier, Janika Raynes, Preston Gibson, Karen Malone, Sarah Davis, Olivia Doak, Judy King, Dayna Wyatt, and the study participants.

---

**Supplemental Table 1.** Characteristics of influenza positive contacts by season, United States, 2017-2023.

| Characteristic | Overall, N = 426<sup>1</sup> | 2017-2018, N = 45<sup>1</sup> | 2018-2019, N = 107<sup>1</sup> | 2019-2020, N = 152<sup>1</sup> | 2021-2022, N = 45<sup>1</sup> | 2022-2023, N = 77<sup>1</sup> |
| --- | --- | --- | --- | --- | --- | --- |
| State |  |  |  |  |  |  |
| TN | 231 (54%) | 24 (53%) | 47 (44%) | 46 (30%) | 45 (100%) | 69 (90%) |
| WI | 195 (46%) | 21 (47%) | 60 (56%) | 106 (70%) | 0 (0%) | 8 (10%) |
| Sex |  |  |  |  |  |  |
| Female | 214 (50%) | 26 (58%) | 52 (49%) | 69 (45%) | 29 (64%) | 38 (49%) |
| Male | 212 (50%) | 19 (42%) | 55 (51%) | 83 (55%) | 16 (36%) | 39 (51%) |
| Race, Ethnicity |  |  |  |  |  |  |
| Asian, Non-Hispanic | 11 (2.6%) | 0 (0%) | 3 (2.8%) | 8 (5.3%) | 0 (0%) | 0 (0%) |
| Black, Non-Hispanic | 20 (4.7%) | 2 (4.4%) | 4 (3.7%) | 6 (3.9%) | 7 (16%) | 1 (1.3%) |
| Hispanic/Latino | 69 (16%) | 6 (13%) | 12 (11%) | 13 (8.6%) | 16 (36%) | 22 (29%) |
| Multiple race, Non-Hispanic | 9 (2.1%) | 0 (0%) | 1 (0.9%) | 6 (3.9%) | 2 (4.4%) | 0 (0%) |
| NH/OPI, Non-Hispanic | 1 (0.2%) | 0 (0%) | 0 (0%) | 0 (0%) | 0 (0%) | 1 (1.3%) |
| Unknown/Refused | 3 (0.7%) | 0 (0%) | 0 (0%) | 0 (0%) | 0 (0%) | 3 (3.9%) |
| White, Non-Hispanic | 313 (73%) | 37 (82%) | 87 (81%) | 119 (78%) | 20 (44%) | 50 (65%) |
| Vaccination Status |  |  |  |  |  |  |
| Unvaccinated | 257 (60%) | 26 (58%) | 50 (47%) | 100 (66%) | 25 (56%) | 56 (73%) |
| Vaccinated | 169 (40%) | 19 (42%) | 57 (53%) | 52 (34%) | 20 (44%) | 21 (27%) |
| Age Group |  |  |  |  |  |  |
| 0-17 | 208 (49%) | 16 (36%) | 60 (56%) | 90 (59%) | 15 (33%) | 27 (35%) |
| 18-49 | 161 (38%) | 19 (42%) | 33 (31%) | 50 (33%) | 18 (40%) | 41 (53%) |
| 50+ | 57 (13%) | 10 (22%) | 14 (13%) | 12 (7.9%) | 12 (27%) | 9 (12%) |
| Flu Type |  |  |  |  |  |  |
| A, H1 | 152 (36%) | 8 (18%) | 41 (38%) | 87 (57%) | 2 (4.4%) | 14 (18%) |
| A, H3 | 139 (33%) | 24 (53%) | 61 (57%) | 0 (0%) | 29 (64%) | 25 (32%) |
| A, no subtyping | 44 (10%) | 1 (2.2%) | 3 (2.8%) | 1 (0.7%) | 14 (31%) | 25 (32%) |
| B, no subtyping | 82 (19%) | 11 (24%) | 1 (0.9%) | 62 (41%) | 0 (0%) | 8 (10%) |
| Unknown | 9 (2.1%) | 1 (2.2%) | 1 (0.9%) | 2 (1.3%) | 0 (0%) | 5 (6.5%) |

<sup>1</sup>n (%)

**Supplemental Table 2.** Characteristics of the cohort & frequencies of asymptomatic infections with follow-up restricted to 5 days^+^, United States, 2017-2023.

|  | Influenza positive contacts | Asymptomatic influenza positive contacts | Modeled Asymptomatic infections* | Contacts without ARI | Modeled Infections without ARI* | Contacts without ILI | Modeled Infections without ILI* |
| --- | --- | --- | --- | --- | --- | --- | --- |
|  | N | N | % (CI) | N | % (CI) | N | % (CI) |
| **Age Group** |  |  |  |  |  |  |  |
| 0 – 17 years | 204 | 11 | 7 (3-12) | 24 | 15 (9-22) | 51 | 29 (22-38) |
| 18 – 49 years | 159 | 19 | 12 (7-20) | 27 | 17 (11-26) | 79 | 53 (44-62) |
| 50+ years | 57 | 4 | 5 (2-15) | 10 | 13 (7-23) | 28 | 48 (35-61) |
| **Season** |  |  |  |  |  |  |  |
| 2017 – 2018 | 42 | 3 | 6 (2-18) | 5 | 11 (5-25) | 15 | 35 (21-52) |
| 2018 – 2019 | 106 | 5 | 4 (2-9) | 11 | 9 (4-17) | 34 | 34 (25-45) |
| 2019 – 2020 | 150 | 5 | 3 (1-9) | 8 | 5 (2-12) | 43 | 34 (25-43) |
| 2021 – 2022 | 45 | 7 | 15 (7-30) | 16 | 36 (22-53) | 26 | 59 (40-75) |
| 2022 – 2023 | 77 | 14 | 17 (9-31) | 21 | 30 (18-46) | 40 | 55 (41-67) |
| **Influenza Vaccination Status** |  |  |  |  |  |  |  |
| Vaccinated | 167 | 18 | 11 (7-17) | 36 | 24 (17-33) | 75 | 51 (41-60) |
| Unvaccinated | 253 | 16 | 5 (3-9) | 25 | 9 (6-14) | 83 | 36 (29-44) |

\*A robust logistic regression model accounting for household clustering was conducted to estimate the proportion of influenza positive household contacts that were asymptomatic, without ARI, and without ILI. Adjusted marginal estimates and 95% confidence intervals are shown as percentages. Age group was adjusted for season and vaccination status. Season was adjusted for age group and vaccination status. Vaccination status was adjusted for age group and season.
<sup>+</sup>Follow-up was restricted to 5 days of follow-up which was the minimum collected per site protocol as sensitivity analyses.

